# Vigorous exercise associates with the abundance of gut bacterial species reflecting energy pathways: an epidemiological cross-sectional analysis within the Lifelines Dutch Microbiome Project

**DOI:** 10.1101/2024.11.05.24316744

**Authors:** Jordi Morwani-Mangnani, Quinten R. Ducarmon, Georg Zeller, Joris Deelen, Marian Beekman, P. Eline Slagboom

## Abstract

**Background:** Regular physical activity (PA) is vital for proper organ functions including the gut. Despite existing research, it remains unclear how the gut microbiome is affected by different intensities of PA, and how other lifestyle factors influence this relationship. Here we study the relation between PA intensity and gut microbiome composition and function in a large Lifelines Dutch Microbiome Project dataset.

**Methods:** A cross-sectional design was performed on 5409 adults aged 40 to 60 from the community-based Lifelines Dutch Microbiome Project and from all these participants metagenomic shotgun data was available. Participants were categorized into sedentary (N=2501), moderate (N=1915), and vigorous (N=993) PA groups, based on self-reported activity levels. We investigated association between PA intensity and microbial diversity, bacterial species, and metabolic pathways by multiple regression models sequentially adjusted for the covariates age/sex, BMI, stool consistency and diet quality/alcohol intake.

**Results:** Vigorous PA, but not moderate PA, was significantly associated with higher gut microbiome alpha diversity (i.e., species richness, Shannon diversity, and Simpson diversity) as compared to sedentary PA. Compared to the sedentary group, the vigorous PA group showed a lower abundance of the bacterial species *Lawsonibacter asaccharolyticus* (β = −0.003, p = 0.042), even after extensive covariate adjustments and correction for multiple testing. Other species were initially also significantly associated with vigorous PA, but they disappeared after adjusting for covariates resulting in a loss of significance. Pathway analysis showed significant enrichment of two distinct metabolic pathways related to cellular energy recycling (*Pyruvate fermentation to acetate and lactate II,* β = 8.11×10^−05^, p = 0.035) and purine metabolism (*Purine ribonucleosides degradation*, β = 3.36×10^−04^, p = 0.039) in participants engaging in vigorous PA as compared to sedentary PA.

**Conclusions:** Vigorous PA is associated with higher gut microbiome diversity and with specific alterations of microbial composition. The lower abundance of *Lawsonibacter asaccharolyticus* within the vigorous PA group may be linked to increased gut permeability. The identified enrichment of microbial fermentation and purine metabolism in vigorous PA hints at a potential role of PA in affecting gut microbiome functionality and host health. The results of our modeling strategy highlight the importance of adjusting for dietary covariates to understand how PA may impact the gut microbiome independently from other influences.

## INTRODUCTION

Physical activity (PA) provides a range of benefits including better cardiovascular health, improved physical functioning, and enhanced mental health (1–5). PA exerts its health-promoting functions on e.g., lung capacity, circulation, muscle mass, and strength. Furthermore, PA likely influences the gut microbiome, a diverse community of microorganisms crucial for digestion, nutrient metabolism, immune function, and intercellular communication with the central nervous system. (6–9). Given the important role of the gut microbiome in host health, understanding how PA modulates can offer valuable insights for optimizing health and preventing disease.

A healthy gut microbiome may be defined by characteristics such as high microbial diversity and a large relative abundance of commensal bacteria. However, more quantitative definitions are still subject to intense research. Current research supports the notion that PA promotes a healthy gut microbiome (6,10). It has, however, not been thoroughly investigated which type of PA changes the relative abundance of specific bacterial species and microbial pathways. Given that the WHO guidelines recommend at least 150 minutes of moderate activity per week and at least 75 minutes of vigorous activity per week, studying the impact of meeting these criteria on gut microbiome is particularly relevant. These levels include sedentary (not meeting guidelines), moderate activity (at least 150 minutes of moderate activity per week), and vigorous activity (at least 75 minutes of vigorous activity per week) (11–14). Noteworthily, previous studies in this area often do not extensively adjust for lifestyle covariates in the relationship between PA and gut microbial health, underscoring the need for a more nuanced exploration of this relationship.

Beyond PA, an array of factors influences gut microbial composition and function. For example, plant-based diets rich in fibers nourish specific commensal bacteria and enhance the production of host-health-promoting metabolites such as short-chain fatty acids (15–17). Similarly, healthier stool consistency, reflecting optimal gut transit time and microbial activity, correlates with a more balanced and diverse microbiome (18). Conversely, factors such as older age and higher BMI, alcohol intake, and medication use are associated with a decline in gut microbial diversity and gut health (19–21). Age-related gut microbiome changes are associated with immunosenescence and inflammaging – low-grade chronic inflammation leading to defective innate immune function, both of which contribute to age-related diseases (19,22). In addition, higher BMI has been associated with less diverse microbiota and with profiles linked to obesity-related risks (20,23). Alcohol intake disrupts microbial communities, compromising intestinal health and immune function (21). While biological sex differences have minimal impact on the gut microbial profiles, differences across genders in lifestyle and behaviors may still modulate gut microbial health (20,24). Given these varied influences on the gut microbiome, it is imperative to understand how PA may impact gut microbial health independently of these known covariates.

The present study aimed to understand the relationship between different intensity levels of PA and the gut microbiome, specifically via microbial alpha diversity, taxonomic species abundance, and microbial pathway abundance. We sequentially accounted for covariates including age, sex, BMI, Bristol stool scale, sequencing read depth, stool consistency, Lifelines Diet Score, daily caloric intake, and alcohol intake. By leveraging the globally unique dataset of the Lifelines Dutch Microbiome Project (DMP), we focused on how higher PA intensities and known covariates influence human health through its modulation of gut microbial composition and function.

## METHODS

### STUDY DESIGN

The DMP cohort expands the Lifelines cohort study, a comprehensive and long-term population-based research initiative (25). Lifelines utilizes a distinctive three-generation approach to investigate the health and health-related behaviors of 167,729 residents of the Netherlands. This study encompasses a wide range of factors that influence health and disease. The Lifelines Cohort Study is conducted according to the principles of the Declaration of Helsinki and per research code UMCG and is approved by the medical ethical committee of the University Medical Center Groningen, The Netherlands.

For the DMP sub-cohort of the Lifelines cohort study, fresh-frozen fecal and blood samples were obtained from Lifelines participants between 2015 and 2016. Whole-genome shotgun sequencing was conducted on portions of 8208 fecal samples for further analysis (26). Participant metadata was grouped into anthropometrics, dietary habits, PA behaviors, diseases, and medication use (27).

### STUDY PROCEDURES

#### Fecal sample collection, DNA extraction, and sequencing

Participants collected and froze fecal samples at home within 15 minutes. Lifelines staff transported these samples on dry ice to a biorepository, where they were stored at −80°C. DNA was extracted using the QIAamp Fast DNA Stool Mini Kit (Qiagen) and QIAcube (Qiagen). Sequencing was done at Novogene, China, on the Illumina HiSeq 2000 platform, generating around 8 Gb of 150 bp paired-end reads per sample (26).

#### Profiling microbiome composition and function

Metagenomes were analyzed as previously described (26). KneadData tools (v0.5.1) trimmed reads to PHRED quality 30 and removed Illumina adapters; Bowtie2 (v2.3.4.1) removed human genome-aligned reads. Taxonomic composition was assessed using MetaPhlAn2 (v2.7.2), and microbial pathways were profiled with HUMAnN2 (v0.11.1) (28).

#### Measurement of Bacterial species and functional pathways

A total of 8208 participants were initially enrolled, from which 5409 individuals (2223 men and 3186 women) between the ages of 20 and 83 year that did not use antibiotics three months prior to stool provision had a complete dataset (FIGURE 1). Stool samples were collected for metagenomic sequencing, analyzing 167 bacterial species and 278 functional pathways. Abundance data was logarithmically transformed to base 10 (Log10), with a pseudocount added as needed. Further details on the protocol are specified in a previous study (28). The MetaCyc encyclopedic database was employed to identify metabolic pathways and enzymes, identifying organism-level specific pathways (29).

**FIGURE 1.**
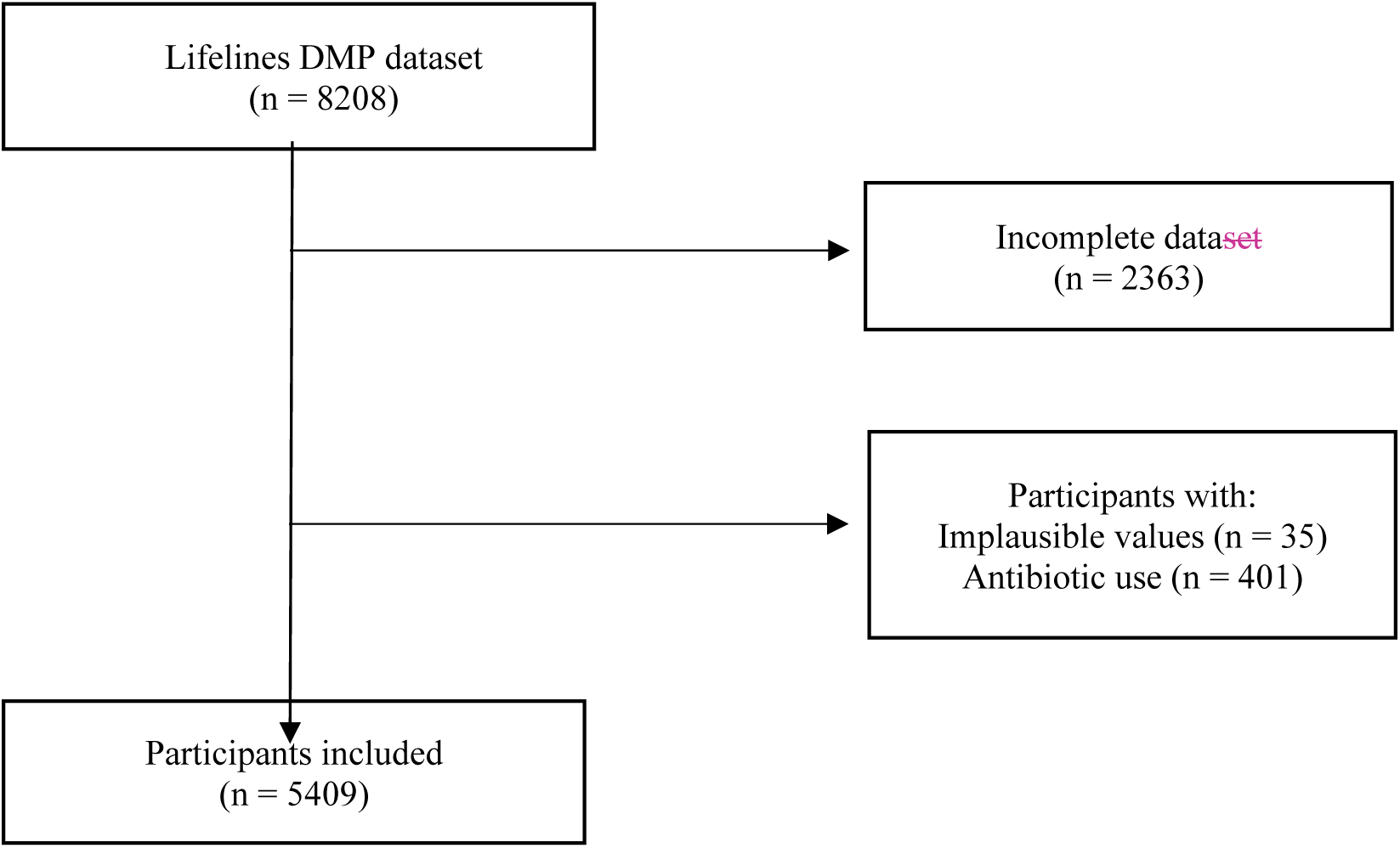
Flow diagram of the final included participants from the original Lifelines DMP cohort.

#### Variables of interest

PA levels were assessed using the widely utilized SQUASH questionnaire, which is recognized nationally for its comprehensive evaluation of PA in the Dutch population. Participants were categorized as sedentary (not meeting WHO guidelines of at least 150 minutes of moderate activity or 75 minutes of vigorous activity per week), moderately active (meeting WHO moderate activity recommendations and engaging more minutes per week in moderate activity than vigorous activity), or vigorously active (meeting WHO vigorous activity recommendations and engaging more minutes per week in vigorous activity than moderate activity).

Demographic variables (i.e., age and sex), traditional covariates (i.e., BMI), gut-related covariates (i.e. Bristol Stool Scale mean taken as the average seven-day score before stool provision, and sequencing read depth), and dietary variables (alcohol intake, Lifelines Diet Score as a proxy of healthiness of a diet, caloric intake) were obtained through questionnaires and physical measurements. More details about these variables are provided in TABLE S1.

### STATISTICAL ANALYSIS

To investigate the associations between PA and microbiome diversity, we conducted multiple regression analyses with microbiome alpha diversity metrics (species richness, Shannon index, Simpson index) as the outcomes, and PA levels (moderate and vigorous compared to sedentary) as the determinants. These analyses were adjusted for confounding factors including age, sex, BMI, Bristol stool scale, sequencing read depth, stool consistency, Lifelines Diet Score, daily caloric intake, and alcohol intake. Additionally, we explored the effects of vigorous PA on bacterial species abundance and functional pathways through sequential adjustments in multiple linear regression models. These models assessed abundance/pathways as outcomes with vigorous PA as the determinant, starting with an unadjusted model (Model 0), followed by adjustments for age and sex (Model 1), BMI (Model 2), Bristol stool consistency (Model 3), Lifelines Diet Score and caloric intake (Model 4), and alcohol intake (Model 5). Given the study’s exploratory nature, False-Discovery Rate (FDR) correction was applied at significance thresholds of p < 0.05 and p < 0.10 for species and functional pathway abundance. All statistical analyses were conducted using R version 4.2.4 and RStudio version 2023.12.1 using the “vegan” package for gut microbiome-specific analyses.

## RESULTS

### BASELINE CHARACTERISTICS

The present study is based on PA levels and gut metagenomics data measured in the Lifelines DMP sub-cohort (26). The study categorized participants based on their PA levels into three groups according to WHO recommendations: sedentary (N=2501), moderate (N=1915), and vigorous (N=993), totaling 5409 individuals with a complete data set and no antibiotic use (FIGURE 1, TABLE 1). PA levels were a defining characteristic of the groups which varied in moderate and vigorous activity. The sedentary group averaged 0.27 ± 0.66 hours per week of vigorous activity and 0.92 ± 0.89 hours of moderate activity. The moderate activity group averaged 0.98 ± 1.92 hours per week of vigorous activity and 11.61 ± 11.35 hours of moderate activity. In contrast, the vigorous activity group reported a significantly higher average of 4.92 ± 3.31 hours per week of vigorous activity and 1.25 ± 1.64 hours of moderate activity. This significant variation in PA levels (p < 2.2×10^−16^) highlights the defining characteristics of each group, with the vigorous activity group showing a markedly higher level of engagement in vigorous activity compared to both the sedentary and moderate activity groups.

**TABLE 1.**
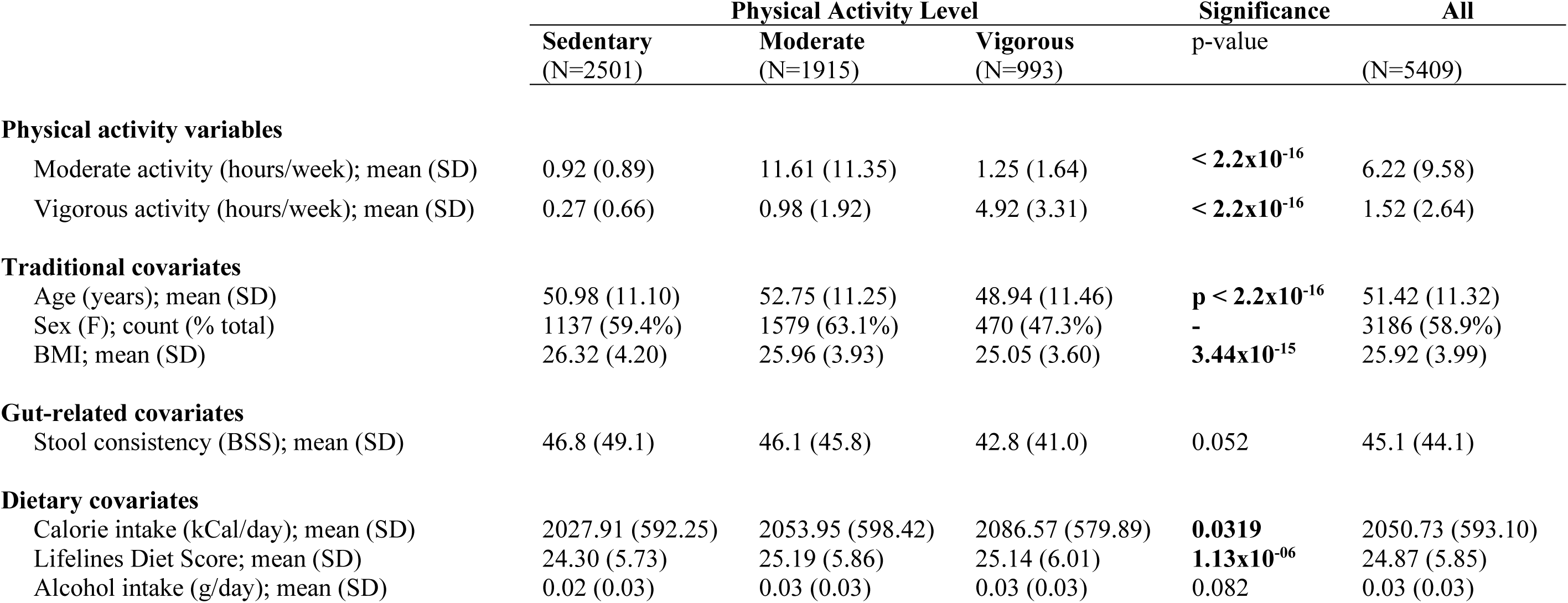
Baseline characteristics of the selected Lifelines DMP cohort participants for analysis. One-way ANOVA tests were carried out to determine overall differences in means across three levels of PA.

|  | Physical Activity Level |  |  | Significance | All |
| --- | --- | --- | --- | --- | --- |
|  | Sedentary<br>(N=2501) | Moderate<br>(N=1915) | Vigorous<br>(N=993) | p-value | (N=5409) |
| **Physical activity variables** |  |  |  |  |  |
| Moderate activity (hours/week); mean (SD) | 0.92 (0.89) | 11.61 (11.35) | 1.25 (1.64) | $< 2.2 \times 10^{-16}$ | 6.22 (9.58) |
| Vigorous activity (hours/week); mean (SD) | 0.27 (0.66) | 0.98 (1.92) | 4.92 (3.31) | $< 2.2 \times 10^{-16}$ | 1.52 (2.64) |
| **Traditional covariates** |  |  |  |  |  |
| Age (years); mean (SD) | 50.98 (11.10) | 52.75 (11.25) | 48.94 (11.46) | $p < 2.2 \times 10^{-16}$ | 51.42 (11.32) |
| Sex (F); count (% total) | 1137 (59.4%) | 1579 (63.1%) | 470 (47.3%) | - | 3186 (58.9%) |
| BMI; mean (SD) | 26.32 (4.20) | 25.96 (3.93) | 25.05 (3.60) | $3.44 \times 10^{-15}$ | 25.92 (3.99) |
| **Gut-related covariates** |  |  |  |  |  |
| Stool consistency (BSS); mean (SD) | 46.8 (49.1) | 46.1 (45.8) | 42.8 (41.0) | 0.052 | 45.1 (44.1) |
| **Dietary covariates** |  |  |  |  |  |
| Calorie intake (kCal/day); mean (SD) | 2027.91 (592.25) | 2053.95 (598.42) | 2086.57 (579.89) | **0.0319** | 2050.73 (593.10) |
| Lifelines Diet Score; mean (SD) | 24.30 (5.73) | 25.19 (5.86) | 25.14 (6.01) | $1.13 \times 10^{-06}$ | 24.87 (5.85) |
| Alcohol intake (g/day); mean (SD) | 0.02 (0.03) | 0.03 (0.03) | 0.03 (0.03) | 0.082 | 0.03 (0.03) |

The analysis of PA levels revealed distinct patterns among the groups. The sedentary group, with an average age of 50.98 ± 11.10 years and the highest BMI of 26.32 ± 4.20, had the lowest calorie intake (2027.91 ± 592.25 kCal/day) and the lowest diet quality, as indicated by the Lifelines Diet Score (24.30 ± 5.73) (TABLE 1)). In contrast, the moderate activity group, which was older 52.75 ± 11.25 years and a had a BMI of 25.96 ± 3.93, had higher caloric intake (2053.95 ± 598.42 kCal/day) and better diet quality (Lifelines Diet Score 25.19 ± 5.86). The vigorous activity group, being the youngest with 48.94 ± 11.46 years and having the lowest BMI of 25.05 ± 3.60, reported the highest calorie intake (2086.57 ± 579.89 kCal/day) and the highest diet quality (Lifelines Diet Score 25.14 ± 6.01). Significant differences were observed across PA levels for age, BMI, calorie intake, and diet quality (all p < 0.05), with no significant variation in stool consistency (p = 0.0923) or alcohol intake (p = 0.0823).

### VIGOROUS PA IS ASSOCIATED WITH HIGHER GUT MICROBIAL ALPHA DIVERSITY

To explore the relationship between gut microbial diversity and PA levels, we assessed these associations in a linear model accounting for various factors, including age, sex, BMI, Bristol Mean, calorie intake, Lifelines Diet Score, and alcohol intake (corresponding to Model 5 in the Methods section). The results, depicted in TABLE 2, indicate that only vigorous activity was significantly associated with higher alpha diversity indices. Specifically, vigorous activity was positively correlated with species richness (β = 3.20, p = 7.80×10^−5^), Shannon diversity (β = 0.0320, p = 0.015), and Simpson diversity (β = 0.002, p = 0.024), even after correction for the full set of covariates (TABLES S2, S3, S4, respectively). In contrast, moderate activity exhibited twice the effect size for these diversity parameters, but none of these associations reached statistical significance.

**TABLE 2.**
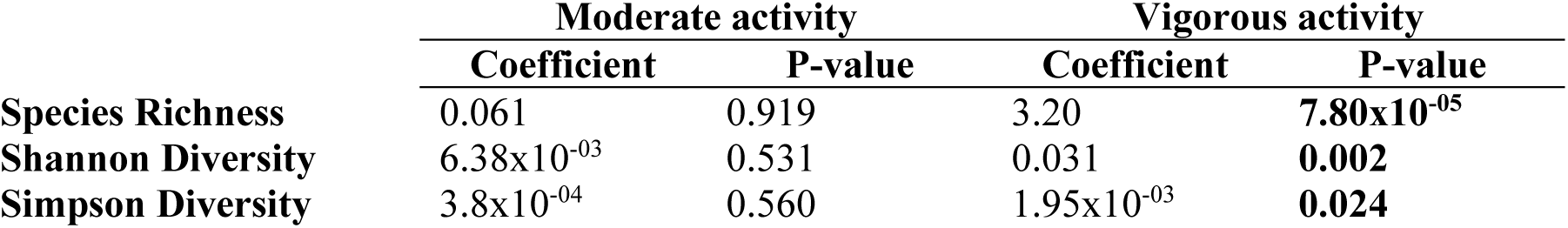
Association of species richness, Shannon Diversity and Simpson Diversity with being moderately active and vigorously active (Model 5 adjusted for age, sex, BMI, Bristol Mean, calorie intake, Lifelines Diet Score, and alcohol intake) Species richness is additionally adjusted for sequencing depth. For results from all models, refer to TABLES S2-S4.

|  | **Moderate activity** |  | **Vigorous activity** |  |
| --- | --- | --- | --- | --- |
|  | **Coefficient** | **P-value** | **Coefficient** | **P-value** |
| **Species Richness** | 0.061 | 0.919 | 3.20 | **7.80x10<sup>-05</sup>** |
| **Shannon Diversity** | 6.38x10<sup>-03</sup> | 0.531 | 0.031 | **0.002** |
| **Simpson Diversity** | 3.8x10<sup>-04</sup> | 0.560 | 1.95x10<sup>-03</sup> | **0.024** |

### VIGOROUS PA IS ASSOCIATED WITH BACTERIAL SPECIES RELATIVE ABUNDANCE

We next examined the associations between the relative abundance of gut microbial species and the levels of vigorous PA compared to the sedentary group. We employed multiple regression models to explore the effects of the potential confounders mentioned above in five models (outlined in the Methods section). The analysis identified several bacterial species that showed significant associations with vigorous PA after adjusting for FDR at 0.05 and 0.1 (see FIGURE 2, TABLE S5, FIGURE S1). In FIGURE S1, we present six sequential models adjusting for age, sex, BMI, Bristol stool consistency, Lifelines Diet Score and caloric intake, and alcohol intake. We present the most interesting transitions in FIGURE 2 of these six models, specifically Models 1, 2, and 5. This stepwise approach reveals how each factor influences the relationship, providing a more nuanced and robust analysis than a single plot or analysis could offer.

**FIGURE 2.**
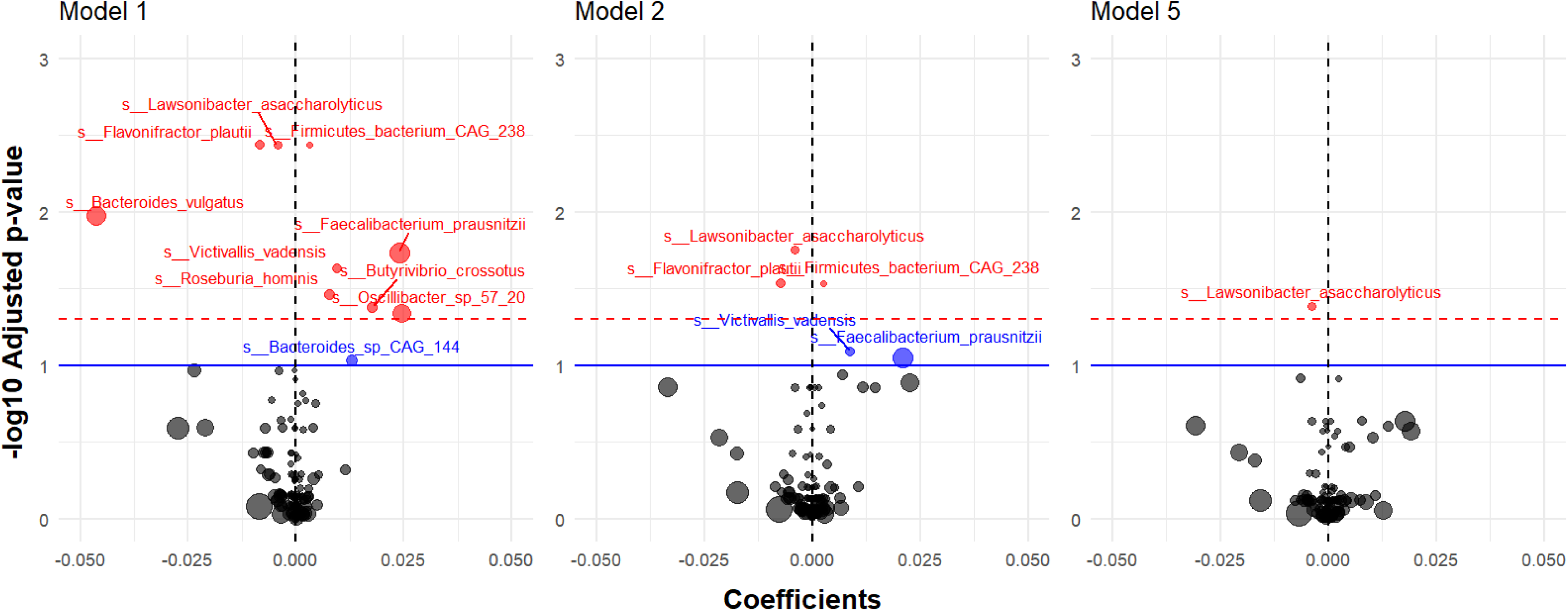
Volcano plots depicting the association between vigorous physical activity levels and gut bacterial species. Model 1 is adjusted for age and sex. Model 2 is additionally adjusted for BMI. Model 5 is additionally adjusted for Bristol Stool Mean, Lifelines Diet Score, and alcohol intake. Significant FDR adjusted p-values below 0.05 are marked in red. Below 0.10 are marked in blue. Higher mean abundances of the species are depicted by larger circumference of the data points.

In the unadjusted model (Model 0), vigorous PA was significantly associated with relative abundances of several bacterial species (TABLE S5, FIGURE S1). *Lawsonibacter asaccharolyticus* and *Flavonifractor plautii* exhibited a significantly reduced relative abundance (β = −4.33×10^−3^, p = 4.29×10^−3^; β = −7.78×10^−3^, p = 0.014, respectively). *Faecalibacterium prausnitzii*, *Oscillibacter sp. 57_20*, and *Bacteroides vulgatus* also showed significantly lower relative abundance with vigorous PA (β = 0.02, p = 0.028; β = 0.024, p = 0.032; β = −0.037, p = 0.045, respectively). In contrast, *Firmicutes bacterium CAG 238* and *Victivallis vadensis* displayed a higher relative abundance with vigorous PA (β = 2.69×10^−3^, p = 0.017; β = 9.23×10^−3^, p = 0.034, respectively).

After adjusting for age and sex (Model 1), the associations between vigorous PA and the gut microbiome remained largely unchanged, with *Roseburia hominis* and *Butyrivibrio crossotus* showing higher relative abundances (β = 7.67×10^−3^, p = 0.034; β = 0.017, p = 0.041, respectively). Further adjustments for BMI (Model 2) reduced the number of significant associations, with only *Lawsonibacter asaccharolyticus*, *Flavonifractor plautii*, and *Firmicutes bacterium CAG 238* maintaining their associations. Adjusting for stool consistency (Model 3) narrowed these down further, leaving only Firmicutes bacterium CAG 238 with a higher abundance and Lawsonibacter asaccharolyticus and Flavonifractor plautii with lower abundances. Finally, in Models 4 and 5, which adjusted for diet quality and alcohol intake, only Lawsonibacter asaccharolyticus remained significantly associated with vigorous PA. Replicating the analysis for moderate PA revealed no significant associations (TABLE S7).

### VIGOROUS PA IS ASSOCIATED WITH REDUCTION AND ENRICHMENT OF MICROBIAL FUNCTIONALITY

Beyond relative abundance of bacterial species, it is important to understand the functional pathways that may further elucidate the metabolic profiles of individuals in the vigorous PA group compared to the sedentary group Therefore, we further tested for associations between gut microbial functional pathways quantified from shotgun metagenomes and levels of vigorous PA using various regression models (Models 0-5, as detailed in the Methods section). The analysis uncovered functional pathway abundances significantly linked to vigorous PA following adjustment for FDR at 0.05 and 0.1 (see FIGURE 3, TABLE S6, FIGURE S2).

**FIGURE 3.**
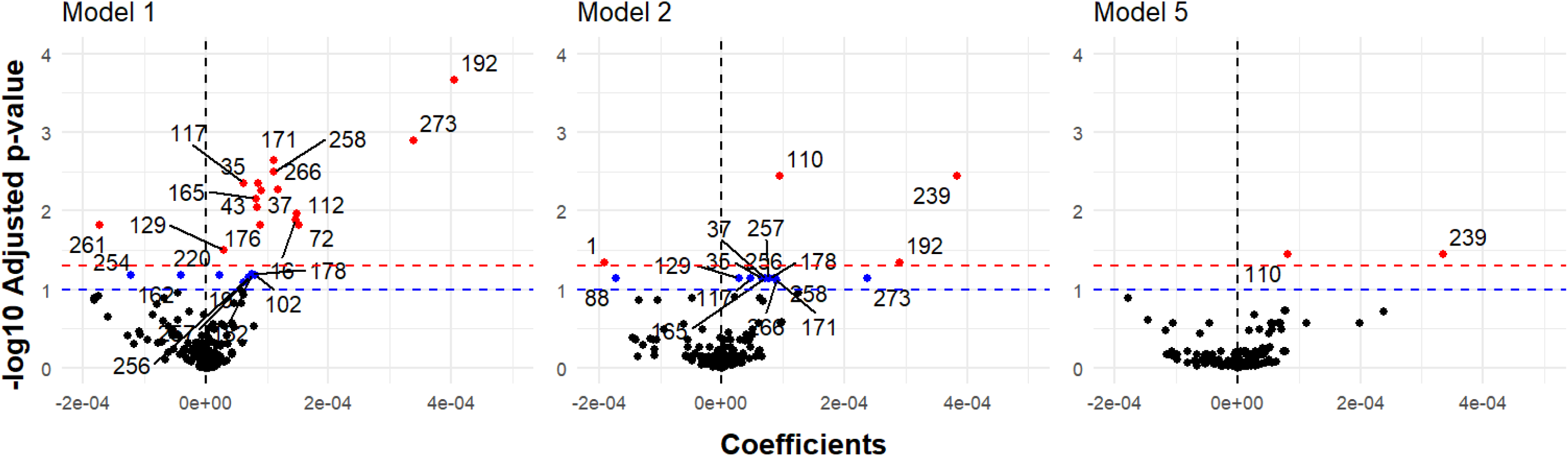
Volcano plots of the multiple linear regression association between vigorous physical activity levels versus sedentary and gut bacterial functional pathways. All 6 Models displayed in FIGURE S5. Model 1 is adjusted for age and sex. Model 2 is additionally adjusted for BMI. Model 5 is additionally adjusted for alcohol intake. Significant FDR adjusted p-values below 0.05 are marked in red. below 0.10 are marked in blue. Legend for coded pathway numbers displayed in table below.

In the unadjusted Model 0, vigorous PA was significantly associated with a range of functional pathways compared to sedentary PA (TABLE S6, FIGURE S2). *Pyruvate fermentation to acetate and lactate II* (hereafter referred to as *pyruvate fermentation)*, *purine ribonucleosides degradation*, and *starch degradation V* showed an enriched microbial functionality (β = 1.06×10^−4^, p = 4.13×10^−4^; β = 4.25×10^−4^, p = 4.13×10^−4^; β = 3.38×10^−4^, p = 5.40×10^−3^, respectively). In contrast, *N10-formyl tetrahydrofolate biosynthesis* and *phosphopantothenate biosynthesis I* exhibited a reduced microbial functionality with vigorous PA (β = 1.22×10^−4^, p = 7.97×10^−6^; β = −1.83×10^−4^, p = 0.026, respectively).

After adjusting for age and sex (Model 1), vigorous physical activity remained consistently associated with pathways related to *pyruvate fermentation*, *purine ribonucleosides degradation*, and *starch degradation V*, *while N10-formyl tetrahydrofolate biosynthesis* and *phosphopantothenate biosynthesis I* showed reduced abundances (FIGURE 3, TABLE S6, FIGURE S2). Adjusting for BMI (Model 2) maintained most associations, though the link with *N10-formyl tetrahydrofolate biosynthesis* and *phosphopantothenate biosynthesis I* weakened. Further adjustments for stool consistency (Model 3) narrowed the associations, with only pyruvate fermentation and purine ribonucleosides degradation pathways remaining enriched. In the final models accounting for diet quality and alcohol intake (Models 4 and 5), only these two pathways persisted. Replicating the analysis for moderate PA revealed no significant associations (TABLE S8).

## DISCUSSION

This study explored the associations of PA on gut microbial diversity, focusing on the relation of vigorous PA and relative species abundance, and function, addressing a key gap in understanding how PA influences gut microbiome characteristics independently from other lifestyle factors in the Lifelines DMP cohort aged 40-60 years. For each of these outcomes we employed multiple regression models by sequentially adding covariates including age and sex, BMI, stool consistence, and diet quality (Models 0 to 5). Only vigorous PA, and not moderate PA, associated with the outcomes. The fully adjusted models revealed that vigorous PA is associated with moderately increase species richness, Shannon diversity, and Simpson diversity indices compared to sedentary PA, independent of all tested covariates. Additionally, *Lawsonibacter asaccharolyticus* was found to have lower relative abundance in those engaging in vigorous PA. Two functional pathways—*Pyruvate fermentation to acetate and lactate II*, and *purine ribonucleosides degradation*—were enriched in the vigorous PA group, even after accounting for confounders. These findings highlight the unique role of vigorous PA in enhancing gut microbial diversity and specific functional pathways, offering new insights into the benefits of PA on gut characteristics.

The stratification based on PA intensity levels is essential to this study. As expected, the vigorous PA group was generally younger, more male-dominated, and had lower BMIs compared to those with moderate or sedentary activity levels, indicating that age and body composition may influence one’s capacity or inclination for intense physical activities. Social, cultural, or physiological factors may influence potential gender differences in activity preferences (30–32). The research also revealed a slight difference in stool consistency with higher PA intensity, indicating a potential physiological effect of PA on gut function (33). Our analyses revealed that vigorous, but not moderate PA, was associated with a higher gut microbial species richness, Shannon diversity, and Simpson diversity. These results indicate that vigorous activity may be associated with the gut microbiome in terms of gut motility, oxygen levels, and metabolic processes (34–37). Additionally, the study observed variations in dietary patterns, with more active individuals consuming slightly more calories and having higher diet quality scores, implying that they may be more health-conscious or have greater nutritional needs (38,39). The study also found that alcohol intake was similar and low across all activity levels.

Our results also potentially suggest that engaging in moderate PA, defined as at least 150 minutes per week of moderate PA, does not exert the same influence on microbial diversity as vigorous PA. This underscores the importance of considering activity intensity levels when studying their effects on the gut microbiome. Longitudinal studies are required to determine whether vigorous PA would indeed be the only class of PA that beneficially affects gut microbial characteristics. This may especially be relevant in recommendations to older populations pressing the relevance of resistance training also for sustaining muscle health.

Our exploration into gut microbial species and functional pathways revealed how vigorous PA, independent from the range of covariates is associated to relative abundance of one bacterial species (*Lawsonibacter asaccharolyticus)* and with higher relative abundance of two functional pathways. *L.asaccharolyticus* was only recently described and, while largely presumed to be a butyrate producer, it essentially remains functionally uncharacterized (40). A reduced abundance of this species has been recently associated with increased gut permeability in stroke patients, which may be associated with detrimental systemic inflammation (41). This may be conceivable given that PA may temporarily induce an inflammatory response following a bout of exercise due to physical stress (37). Furthermore, we found that vigorous PA is associated with higher metagenomic levels of the *Pyruvate fermentation to acetate and lactate II* pathway after extensive covariate adjustments. Acetate and lactate play a crucial role in healthy gut homeostasis; acetate supports intestinal epithelial integrity and serves as an energy source, while lactate plays a role in gluconeogenesis, aiding metabolic balance (42). Similarly, the pathway *Purine ribonucleosides degradation* also displayed higher metagenomic levels in the vigorous PA group after covariate adjustment. This pathway involves the degradation of purine ribonucleosides, crucial for nucleotide recycling and energy metabolism essential for cellular repair and function (43). Enhanced breakdown of purine ribonucleosides may potentially suggest an association with energy requirements, cellular resilience, and metabolic efficiency (44)

Diet quality, as embodied by the Lifelines Diet Score, emerged as a crucial covariate that may be more directly associated with *Flavonifactor plautii* and *N10-formyl tetrahydrofolate biosynthesis* than to vigorous PA. *F. plautii* initially displayed a lower relative abundance with vigorous PA, but this effect diminished once dietary adjustments were applied. This may suggest that this species, which degrades flavonoids may have a stronger connection to dietary factors, particularly fiber and flavonoid intake rather than vigorous PA itself (45). Thus, while vigorous PA might contribute to a healthier gut environment, dietary factors, including fiber and flavonoids, appear to have a more pronounced impact on the abundance of *F. plautii*. Similarly, the adjustment for diet eliminated the association of vigorous PA with the metagenomic levels with *N10-formyl tetrahydrofolate biosynthesis* is involved in the synthesis of N10-formyl tetrahydrofolate, a key compound in one-carbon metabolism and nucleotide synthesis, essential for cell growth and overall metabolism (46). Previous research suggests that the *N10-formyl tetrahydrofolate biosynthesis* has probiotic properties that are associated with the abundance of commensal bacterial species that have beneficial effects on obesity and diabetes (47). Given that folate, the original substrate of the upstream reaction in this metabolic pathway may only be obtained through diet, it is plausible to infer that this covariate is more influential than PA itself.

This study has notable strengths, including the use of a large cohort with metagenomic profiling, which provides high resolution and statistical power for examining the relationship between vigorous physical activity and gut microbiome diversity. The sequential adjustment for covariates such as age, sex, BMI, stool consistency (measured using the 7-day average Bristol Stool Chart), diet quality, and alcohol intake helps clarify whether PA is independently associated with gut microbial species and pathways. Initially, vigorous PA appeared to be associated with certain bacterial species, but these associations often diminished after adjusting for these covariates. This suggests that microbiome changes may not be directly driven by PA itself, but rather influenced by other factors, such as diet, which was assessed using the Lifelines Diet Score. Diet, known for its significant impact on microbial composition, often eclipses the effects of PA as evidenced in this study, while BMI and stool consistency further modulate gut microbial populations by affecting metabolic health and gut transit time, respectively (48–50).

However, several limitations must be acknowledged. The cross-sectional design limits our ability to establish causal relationships between PA and gut microbiome diversity, allowing us only to observe associations. The present study is only based on one study cohort without replication, therefore limiting its generalizability. Additionally, self-reported data on PA and alcohol intake may be susceptible to biases, such as social desirability and recall inaccuracies. Using objective measurements, like accelerometers, could improve the precision of PA data and their associations with bacterial relative abundances and metagenomic pathway levels. Furthermore, many bacterial species within the gut microbiome remain poorly understood in terms of their specific functions, adding complexity to interpreting their roles concerning PA. These factors highlight the need for caution in concluding the direct effects of PA on the gut microbiome.

In conclusion, this study elucidates significant associations between vigorous PA and the abundance of specific gut bacterial species and functional pathways independent of diet and body composition. While moderate PA was not linked to gut microbiome outcomes, vigorous PA showed associations with the abundance of one bacterial species and two metagenomic pathways. These findings suggest that PA alone, even at higher intensities, may not be as effective as diet to modulate the gut microbiome for health benefits. These insights provide an incentive for future research to extensively account for covariates to longitudinally explore whether and how vigorous or in combination with moderate PA may directly or indirectly modulate the gut microbiome. The observed associations underscore the importance of considering PA and covariates in the broader context of gut health and overall well-being.

## Data Availability

All data produced in the present study are available upon reasonable request to the LifeLines team.

## ACKNOWLEDGMENTS

The authors wish to acknowledge the services of the Lifelines Cohort Study, the contributing research centres delivering data to Lifelines, and all the study participants. The Lifelines initiative has been made possible by subsidy from the Dutch Ministry of Health, Welfare and Sport, the Dutch Ministry of Economic Affairs, the University Medical Center Groningen (UMCG), Groningen University and the Provinces in the North of the Netherlands (Drenthe, Friesland, Groningen).

This work is funded by the VOILA Consortium (ZonMw 457001001), which had no role in the design and conduct of the study; collection, management, analysis, and interpretation of the data; and preparation, review, or approval of the manuscript.

## ABBREVIATIONS

PA: Physical Activity
DMP: Dutch Microbiome Project
WHO: World Health Organization
SQUASH: Short QUestionnaire to ASsess Health enhancing physical activity
FFQ: Food Frequency Questionnaire

---

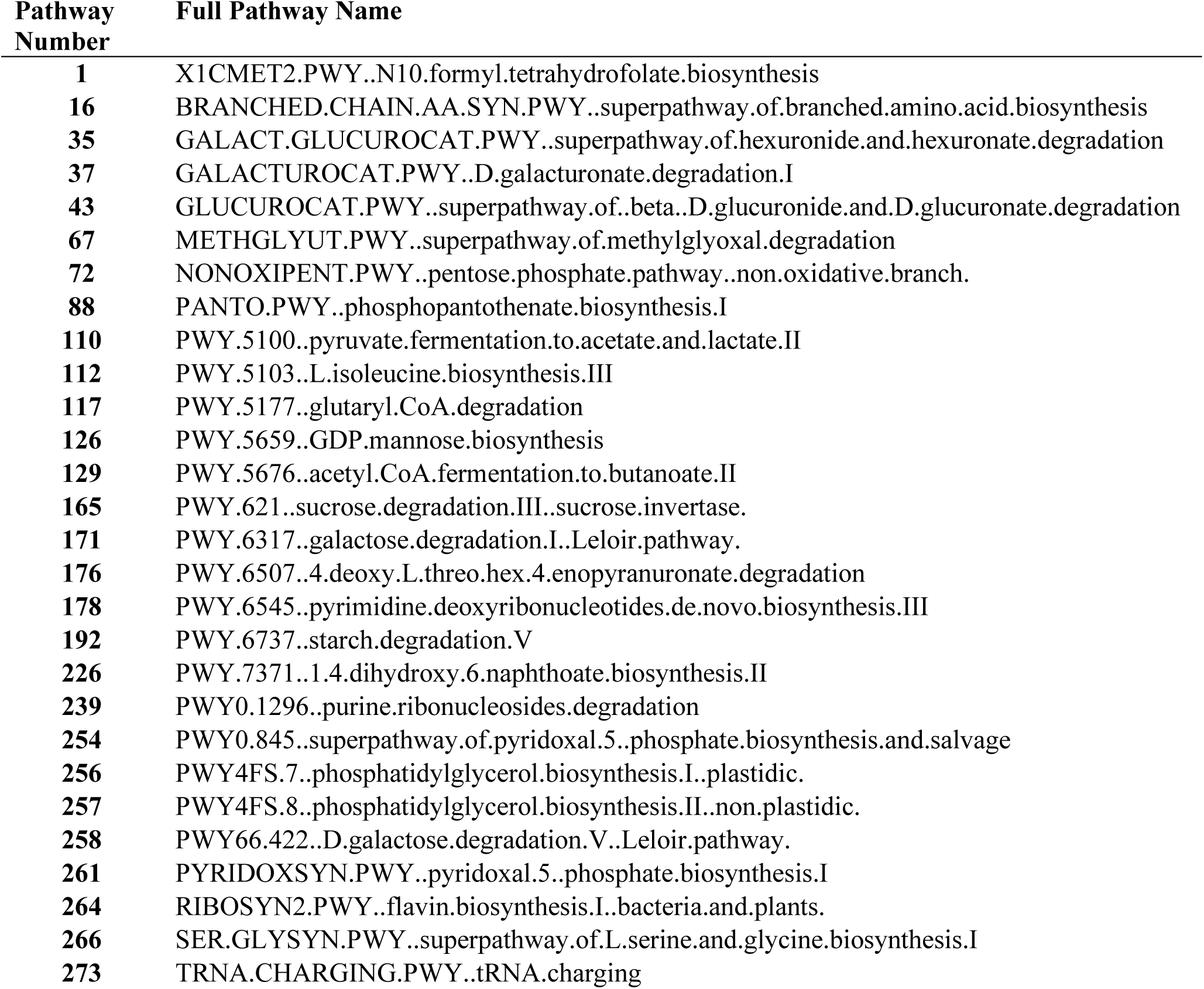

## Notes

### Competing Interest Statement

The authors have declared no competing interest.

### Author Declarations

The Lifelines Cohort Study is conducted according to the principles of the Declaration of Helsinki and per research code UMCG and is approved by the medical ethical committee of the University Medical Center Groningen, The Netherland. All data were available upon request to the LifeLines team in Groningen, the Netherlands.

