## Supplemental Figures 1&2 for "Vigorous exercise associates with the abundance of gut bacterial species reflecting energy pathways: an epidemiological cross-sectional analysis within the Lifelines Dutch Microbiome Project"

**FIGURE S1. Volcano plots of the multiple linear regression association between vigorous physical activity levels versus sedentary and gut bacterial species.** Model 0 is unadjusted. Model 1 is adjusted for age and sex. Model 2 is additionally adjusted for BMI. Model 3 is additionally adjusted for Bristol Mean. Model 4 is additionally adjusted for calorie intake and Lifelines Diet Score. Model 5 is additionally adjusted for alcohol intake. Significant FDR adjusted p-values below 0.05 are marked in red. Below 0.10 are marked in blue. Higher mean abundance of the species depicted by larger circumference of data points.

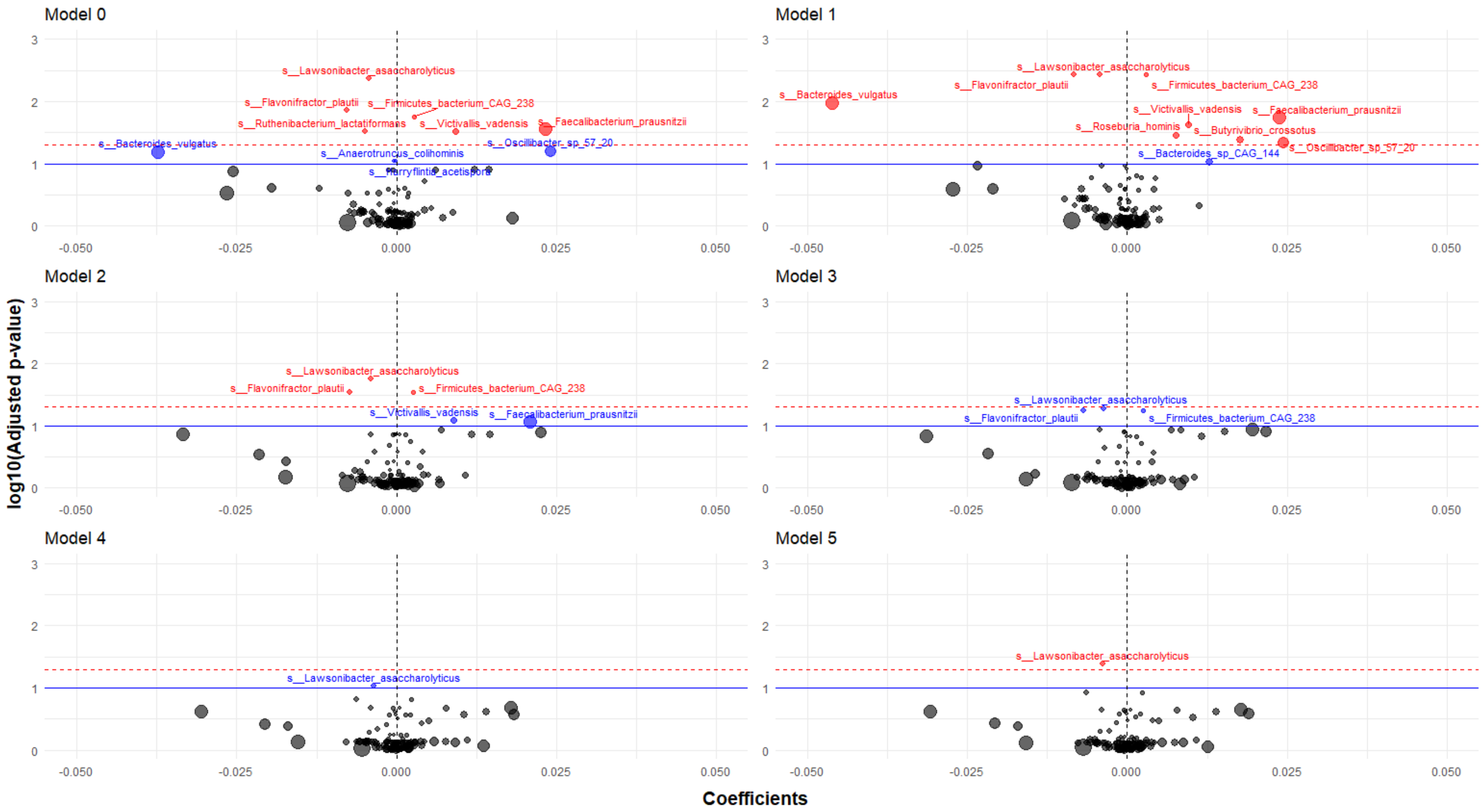

**Figure S2. Volcano plots of the multiple linear regression association between vigorous physical activity levels versus sedentary and gut bacterial functional pathways.** Model 0 is unadjusted. Model 1 is adjusted for age and sex. Model 2 is additionally adjusted for BMI. Model 3 is additionally adjusted for Bristol Mean. Model 4 is additionally adjusted for calorie intake and Lifelines Diet Score. Model 5 is additionally adjusted for alcohol intake. Significant FDR adjusted p-values below 0.05 are marked in red. below 0.10 are marked in blue. Legend for coded pathway numbers displayed in table below.

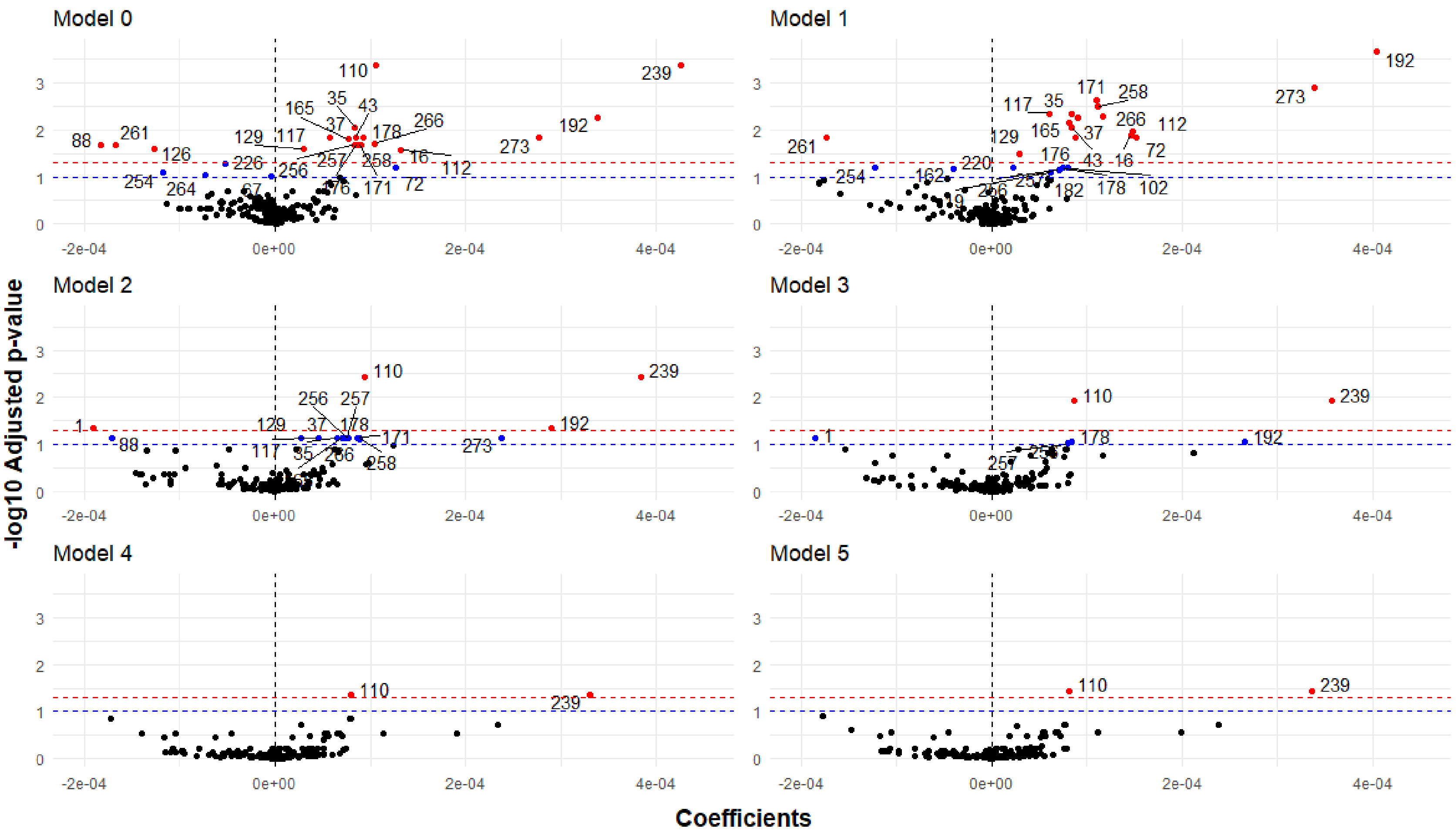

| Pathway Number | Full Pathway Name |
|---|---|
| **1** | X1CMET2.PWY..N10.formyl.tetrahydrofolate.biosynthesis |
| **16** | BRANCHED.CHAIN.AA.SYN.PWY..superpathway.of.branched.amino.acid.biosynthesis |
| **35** | GALACT.GLUCUROCAT.PWY..superpathway.of.hexuronide.and.hexuronate.degradation |
| **37** | GALACTUROCAT.PWY..D.galacturonate.degradation.I |
| **43** | GLUCUROCAT.PWY..superpathway.of..beta..D.glucuronide.and.D.glucuronate.degradation |
| **67** | METHGLYUT.PWY..superpathway.of.methylglyoxal.degradation |
| **72** | NONOXIPENT.PWY..pentose.phosphate.pathway..non.oxidative.branch. |
| **88** | PANTO.PWY..phosphopantothenate.biosynthesis.I |
| **110** | PWY.5100..pyruvate.fermentation.to.acetate.and.lactate.II |
| **112** | PWY.5103..L.isoleucine.biosynthesis.III |
| **117** | PWY.5177..glutaryl.CoA.degradation |
| **126** | PWY.5659..GDP.mannose.biosynthesis |
| **129** | PWY.5676..acetyl.CoA.fermentation.to.butanoate.II |
| **165** | PWY.621..sucrose.degradation.III..sucrose.invertase. |
| **171** | PWY.6317..galactose.degradation.I..Leloir.pathway. |
| **176** | PWY.6507..4.deoxy.L.threo.hex.4.enopyranuronate.degradation |
| **178** | PWY.6545..pyrimidine.deoxyribonucleotides.de.novo.biosynthesis.III |
| **192** | PWY.6737..starch.degradation.V |
| **226** | PWY.7371..1.4.dihydroxy.6.naphthoate.biosynthesis.II |
| **239** | PWY0.1296..purine.ribonucleosides.degradation |
| **254** | PWY0.845..superpathway.of.pyridoxal.5..phosphate.biosynthesis.and.salvage |
| **256** | PWY4FS.7..phosphatidylglycerol.biosynthesis.I..plastidic. |
| **257** | PWY4FS.8..phosphatidylglycerol.biosynthesis.II..non.plastidic. |
| **258** | PWY66.422..D.galactose.degradation.V..Leloir.pathway. |
| **261** | PYRIDOXSYN.PWY..pyridoxal.5..phosphate.biosynthesis.I |
| **264** | RIBOSYN2.PWY..flavin.biosynthesis.I..bacteria.and.plants. |
| **266** | SER.GLYSYN.PWY..superpathway.of.L.serine.and.glycine.biosynthesis.I |
| **273** | TRNA.CHARGING.PWY..tRNA.charging |
